# “It’s a helluva journey”: A qualitative study of patient and clinician experiences of nausea and vomiting syndromes and functional dyspepsia

**DOI:** 10.1101/2022.08.09.22278607

**Authors:** Gabrielle Sebaratnam, Mikaela Law, Elizabeth Broadbent, Armen A Gharibans, Christopher N Andrews, Charlotte Daker, Greg O’Grady, Stefan Calder, Celia Keane

## Abstract

**Background:** Chronic gastroduodenal disorders including chronic nausea and vomiting syndrome, gastroparesis, and functional dyspepsia, are challenging to diagnose and manage. The diagnostic and treatment pathways for these disorders are complex, costly and overlap substantially; however, experiences of this pathway have not been thoroughly investigated. This study therefore aimed to explore clinician and patient perspectives on the current clinical pathway.

**Methods:** Semi-structured interviews were conducted between June 2020 and June 2022 with 11 patients with chronic nausea and vomiting syndrome alone or with functional dyspepsia (based on Rome IV criteria) and nine gastroenterologists who treat these conditions. Interviews were recorded, transcribed, and thematically analyzed using an iterative, inductive approach.

**Results:** Five key patient themes were identified: (1) the impacts of their chronic gastroduodenal symptoms, (2) the complexity of the clinical journey, (3) their interactions with healthcare providers, (4) the need for advocacy, and (5) their experience of treatments. Five key clinician themes were also identified: (1) these conditions were seen as clinically complex, (2) there is an uncertain and variable clinical pathway, (3) the nuance of investigations, (4) these conditions were difficult to therapeutically manage, and (5) there are barriers to developing a therapeutic relationship.

**Conclusions:** Findings indicate that both patients and clinicians are dissatisfied with the current clinical care pathways for nausea and vomiting syndromes and functional dyspepsia. Recommendations included the development of more clinically relevant and discriminant tests, standardization of the diagnostic journey, and the adoption of a multidisciplinary approach to diagnosis and treatment.

## Introduction

Chronic gastroduodenal disorders include nausea and vomiting syndromes (NVS; used here to encompass both chronic nausea and vomiting syndrome (CNVS) and gastroparesis) ^1^ and functional dyspepsia (FD) ^2^. These conditions are challenging to define, diagnose and clinically manage ^3–5^. NVS and FD are highly prevalent, affecting up to 10% of the global population and contributing almost 30% of new patient referrals to gastroenterology clinics ^6,7^. The predominant symptoms in NVS are nausea and vomiting ^1^, whereas FD is the experience of chronic early satiation, excessive fullness, and postprandial fullness ^2^. However, there is significant symptom overlap between CNVS, gastroparesis and FD ^8,9^.

An incomplete understanding of the underlying pathology, heterogeneous symptom presentation, and lack of organic biomarkers for these disorders poses challenges to clinical diagnosis ^2,10–12^. Although a number of international guidelines exist for the management of NVS and FD, these often lack quantitative evidence-based data, resulting in controversy regarding how these conditions are defined and clinically managed ^13,14^. An absence of multidisciplinary approaches and limited availability of pharmacological treatments have been identified as key challenges to the management of these conditions ^15^. To address these difficulties, patients are often extensively investigated to exclude organic pathologies and undergo trial and error of off-label medications and diets to manage symptoms ^2,15,16^. This results in higher healthcare utilization ^17,18^ and more hospital admissions ^19^ compared to other gastroenterology patient groups.

Limited literature has specifically assessed the impacts of these challenges on the clinical care pathways for NVS and FD ^20–22^. Clinical care pathways are tools used to guide the flow of healthcare activities required for the management of a patient’s care for a given health condition ^20,23^. The care pathway refers to the journey that patients experience to access diagnosis and treatment for their symptoms. Anecdotal evidence posits that the clinical care pathway for patients with NVS and FD is convoluted and complex; however, there is little academic research to substantiate these claims ^24^. There is evidence to suggest that patients are dissatisfied with the current clinical care pathway due to a lack of diagnosis and treatment, perceived need for more testing, and frustration with the health system ^25^. Diagnostic uncertainty also appears to contribute to patient anxiety and frustration ^22,26^. However, a thorough understanding of the inadequacies of the current clinical care pathways and the impacts these have on the lived experiences with these conditions is unclear. Previous research corroborates the challenging lived experiences of patients with FGIDs, including; poor health related quality of life, stigma from healthcare providers and social networks, feelings of loss and sadness, and psychological distress ^21,27–29^. As such, efforts to better understand the clinical care pathway for patients with NVS and FD and how the clinical challenges impact their lived experiences is warranted ^30^.

There is also limited literature focusing on clinician experiences of this clinical care pathway ^31^. Clinician perspectives of managing pediatric functional gastrointestinal disorders (FGIDs) highlight the need to develop strong therapeutic relationships with patients and the need to frame management within the biopsychosocial model ^32^. Other findings suggest that primary care physicians lack confidence with diagnosing and managing adult FGID patients ^25^. Further targeted qualitative research is needed to better understand gastrointestinal clinicians’ perspectives of the clinical care pathways for non-paediatric populations and identify how the provision of care can be improved.

This study therefore aimed to qualitatively explore both patient and clinician perspectives on the current clinical care pathways for NVS and FD. Secondary aims included comparing patient and clinician perspectives and identifying areas for improvement.

## Methods

A qualitative interview study was conducted to gather a rich dataset on the experiences of patients and clinicians with the current clinical care pathway for NVS and FD. Ethics approval was granted by the Auckland Health Research Ethics Committee (AH1352). The study was reported as per the Standards for Reporting Qualitative Research (SRQR) ^33^.

### Researcher Reflexivity

The researchers were a group of health psychologists, medical professionals, and bioengineers. Although the interviewers did not have a prior relationship with the patient participants, some of the researchers were known to the clinician participants. This aided recruitment; however, could have introduced bias. Interviews were conducted by a clinical academic who did not have any prior relationships with the study participants. Two health psychology researchers conducted the analysis with oversight from medical professionals and a bioengineer with a specialist interest in gastric electrophysiology. The interpretation of the thematic analysis was informed by these perspectives.

### Sample

Patients with NVS or FD and consultant gastroenterologists who saw and treated patients with these conditions were recruited. Patient participants were recruited from clinical referrals, and online social media advertising, including via patient peer support groups. Maximum diversity sampling was attempted to gain a diverse clinician population. This was based on gender, age, scope of clinical practice (generalist vs motility specialist gastroenterologists), practice type (public/private, regional vs urban centers, academic vs non-academic clinicians), and years in clinical practice. Patients were eligible for the study if they met the diagnostic criteria for CNVS and/or FD (as per Rome IV criteria) ^2^, were aged ≥18 years, and were able to provide informed consent. Individuals who were unable to speak or read English and vulnerable participants (e.g., individuals with a known cognitive impairment and prisoners) were excluded. Data saturation was reached with the sample, with no new themes emerging after the 11th patient participant, and 9th clinician participant. Participant recruitment and data collection was completed between June 2020 and June 2022.

### Protocol

All participants took part in individual semi-structured interviews that were conducted online via a video conferencing platform. All interviews were primarily conducted by CK, who is experienced in qualitative research methods; however, GS assisted with three interviews. Written informed consent was obtained from each participant prior to the start of the interview. Separate patient and clinician interview schedules were developed to guide the interview, with flexibility to explore any other important issues that arose during discussion. These interview schedules were developed based on the aims of the study. Patient interviews explored their symptom experience (including duration, diagnosis, management, and impact on everyday life), their experience of the clinical care pathway (including experiences with healthcare professionals, testing, and treatment), and the impacts of this pathway. Clinician interviews explored their experiences diagnosing and managing patients with NVS and FD. Interviews were recorded, transcribed verbatim, and de-identified prior to analysis.

### Analysis

Iterative, inductive thematic analysis was conducted by two independent coders (GS, ML) ^34^. The two coders individually read through the transcripts to familiarize themselves with the data and then grouped the ideas into common patterns (i.e., themes). Triangulation occurred, with the two coders meeting to discuss and collectively develop a set of key themes with further subthemes, based on their individual analyses. These themes were then reviewed and refined by a wider research panel (CK, CNA, GOG, SC). The transcripts were reviewed again against these developed themes to ensure that no important information had been missed. This process was done separately for the clinician and patient interviews to generate two sets of results.

## Results

### Demographics

In total, 11 patients and nine clinicians were interviewed. All patients met the Rome IV criteria for CNVS; however, ten also met the criteria for FD. During their interviews, eight patients mentioned that they had been diagnosed with gastroparesis, but this was not able to be corroborated clinically as researchers did not access participant medical records. The clinician sample comprised of general gastroenterologists (n = 4) and gastroenterologists specializing in gastrointestinal motility (n = 5). A comprehensive overview of the demographic characteristics of the sample is presented in *Table 1*. The clinician interviews lasted between 17-46 minutes and the patient interviews between 40-122 minutes.

**Table 1.** Demographic and clinical characteristics

| Characteristic | Patients<br>(n= 11) | Clinicians<br>(n= 9) |
| --- | --- | --- |
| Age (years), median (IQR) | 35 (25 - 42) | 42 (39 - 44) |
| Female, n (%) | 11 (100) | 3 (33) |
| Ethnicity, n (%) |  |  |
| Caucasian | 9 (82) | 5 (56) |
| Māori | 1 (9) | 1 (11) |
| Indian | 1 (9) | 0 (0) |
| Chinese | 0 (0) | 1 (11) |
| Other | 0 (0) | 2 (22) |
| Rome IV Diagnosis, n (%) |  |  |
| CNVS only | 1 (9%) | - |
| CNVS and FD | 10 (91%) | - |
| Specialty, n (%) |  |  |
| General gastroenterologist | - | 4 (44) |
| Motility specialist | - | 5 (56) |
| Country of practice, n (%) |  |  |
| Australia | - | 2 (22) |
| Canada | - | 2 (22) |
| New Zealand | - | 5 (56) |
| Practice experience (years), median (IQR) | - | 7 (2 - 10) |
| Public practice only, n (%) | - | 3 (33) |
| Public and private practice, n (%) | - | 6 (67) |
| Regional practice only, n (%) | - | 1(11) |
| University affiliated, n (%) | - | 5 (56) |

Notes: IQR = interquartile range; CNVS = chronic nausea and vomiting syndrome; FD = functional dyspepsia.

### Patient Themes

Five key themes, with several subthemes, were identified from the patient interviews. The hierarchy of the themes is outlined in *Table 2* with example quotations and briefly described in-text. Subthemes have been presented in-text using italics.

**Table 2.**
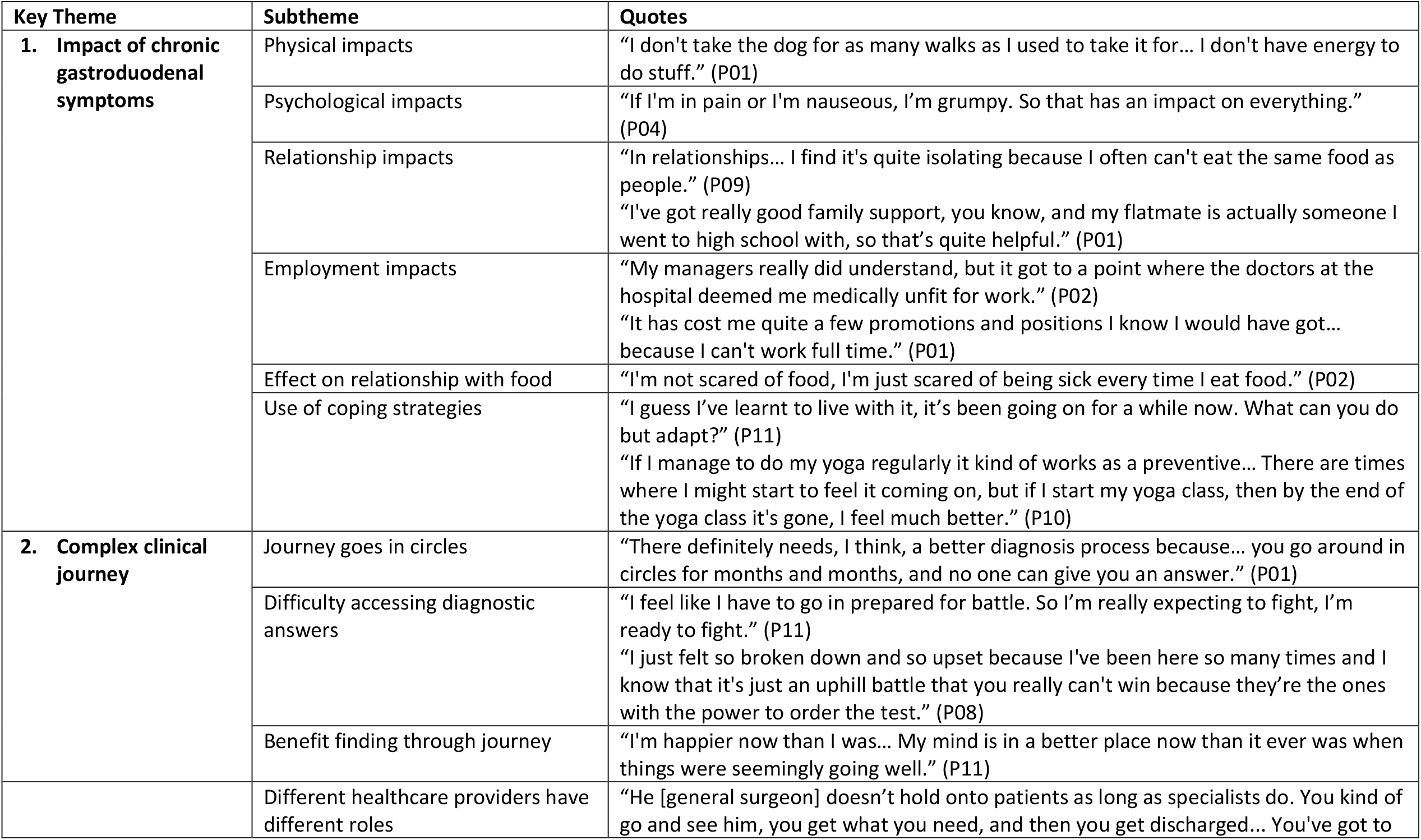
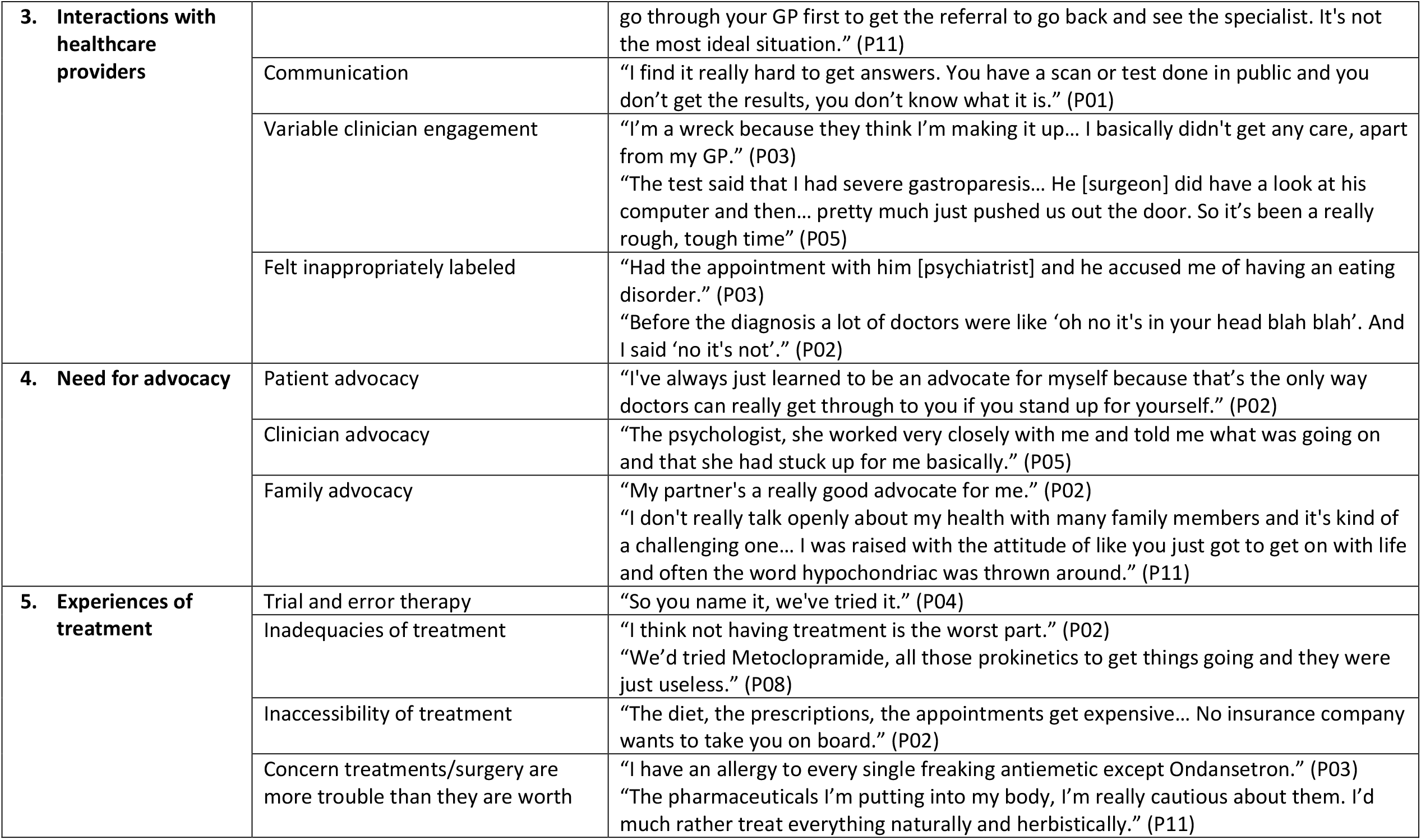
Hierarchy of the themes and subthemes identified from the patient interviews with example quotations

#### Theme 1: Impact of chronic gastroduodenal symptoms

The first patient theme reflected the various impacts that chronic gastroduodenal symptoms had on patients’ lives. Firstly, patients mentioned the *physical impacts* of their symptoms and discussed how the presence of symptoms limited their physical functioning in their day-to-day lives. Symptoms also resulted in *psychological impacts*, by negatively affecting patients’ mood and mental state, and *impacted their relationships with others*. Eating and drinking are central to socialization, which leads to isolation for patients who cannot partake in these activities. Patients discussed feeling like they were a burden on their family and friends. However, their experiences also revealed the strength of their relationships and highlighted the quality of their support system. Patients discussed the *impact that their symptoms had on their employment opportunities*, commenting on the difficulties they had with work and/or study. Lastly, the patients’ symptoms *impacted their relationship with food*, with patients mentioning an avoidance of food and fear of eating.

To minimize these impacts, patients employed *the use of coping strategies*. They didn’t want their disease to control their life, so instead found ways to learn to live with their symptoms and make it work. There was heterogeneity in the coping strategies used, with each patient finding an approach that worked for them (e.g. preparing meals in advance, limiting physical activity, using cannabis, or doing yoga).

#### Theme 2: Complex clinical journey

The second patient theme reflected the complex clinical journey patients experienced while trying to get a diagnosis and treatment for their symptoms. Their journey was not straightforward and instead appeared to *go in circles*. This journey was prolonged by a series of exhaustive tests and long wait times, which resulted in a feeling of not getting anywhere. Patients were also often sent back and forth between different healthcare providers, further convoluting their pathway. Because of this, patients found that they had *difficulty accessing diagnostic answers*. Patients reported that their healthcare providers didn’t always take their concerns seriously. Unfortunately, this resulted in patients viewing seeking healthcare as a battle and often led to an avoidance of accessing healthcare. However, patients did experience *benefit finding during their journey*. Having a diagnosis removed uncertainty and helped narrow down treatment options. The extensive testing, although prolonged, provided reassurance that other medical concerns had been ruled out. Some patients also showed personal growth from their experiences, discussing how they were proud of their body and that their journey had forced self-discovery and learning to find joy.

#### Theme 3: Interactions with healthcare providers

The third key patient theme discussed the extent and quality of the interactions that patients had with their healthcare providers. Throughout their journey, patients had interacted with many *different healthcare providers who played different roles in their care*. Across these providers, patients found that a lack of *communication* was an important determinant for the quality of their interactions. Patients often felt like they were not always told what was happening and it was hard to get answers. In contrast, patients highlighted the importance of having healthcare providers who were supportive and had clear communication. Alongside this, healthcare providers were seen as having *variable engagement*. Although some were seen as helpful and supportive, patients discussed experiences of providers appearing to dismiss their concerns or not take their symptoms seriously. There was concern that this lack of engagement might result in them not being tested or treated for the actual issue. This view stemmed from the fact that patients *felt inappropriately labeled*. This resulted in patients being referred to specialists that were unable to help with their gastrointestinal symptoms (e.g. eating disorder clinics, gynecologists, or psychologists), further convoluting their diagnostic and treatment pathway.

#### Theme 4: Need for advocacy

The fourth key patient theme reflected the need for advocacy in their care. *Patients felt the need to self-advocate* for testing and treatment and believed they had to fight so that their concerns would be taken seriously. Patients also discussed the importance of *clinician advocacy*. Some patients had experience with healthcare providers that had advocated for them in the past and mentioned the importance of this for the progression of their care. In contrast, the absence of clinician advocacy was seen as detrimental. Lastly, the importance of *family advocacy* was discussed. Some patients had family members who advocated for them; however, others experienced unsupportive family members, and felt like they had to self-advocate in their own families.

#### Theme 5: Experiences of treatment

The last key patient theme explored the experiences that patients had with treatment. Throughout their clinical journey, the patients had tried many different treatments, through *trial and error therapy*. Unfortunately, patients experienced many *inadequacies of their treatments*. They found that the majority of treatments were ineffective and in response wanted their disorders to be researched more so that effective treatments could be made available. Patient discussions also reflected the *inaccessibility of treatment options*, as many treatment options were difficult to access due to cost and physical availability. Lastly, there was an overall concern that *treatments were more trouble than they were worth*. Patients mentioned experiences of side-effects and allergies; whilst others told stories about how some treatments, such as feeding tubes, were hard for their body to tolerate. There was also concern over the long-term effects of treatments, especially when these were unknown.

### Clinician Themes

Five key themes, each with a number of subthemes, were also identified from the clinician interviews. The hierarchy of the themes is shown in *Table 3*, with example quotations, and described briefly below.

**Table 3.** Hierarchy of the themes and subthemes identified from the clinician interviews with example quotations

| Key Theme | Subtheme | Quotes |
| --- | --- | --- |
| **1. Clinical complexity** | Viewed as a complex set of disorders | "They're a complex group... So I think because there's that clinical challenge people feel less comfortable with it." (C04)<br>"Often it's a very difficult patient group to treat, and so it does take quite a long time. And I think also that is particularly fueled by an uncertainty from the physician treating as to are they missing something or are they on the right track." (C08) |
|  | Difficulty of diagnosis | "It's [Rome IV criteria] useful for trials, you need to have a standardized assessment, but when it comes to clinical practice, it's not really all that helpful." (C02)<br>"I'm a bit dubious of the reliability of the gastric emptying studies that we get... Our radiologists are doing these studies infrequently and perhaps aren't as expert at interpreting the findings." (C07) |
|  | Need for a multidisciplinary collaborative approach | "[I have suggested] some sort of comprehensive assessment unit analogous to the liver transplant unit; where people go for an assessment, they have a 360 assessment with all the expert assessors able to do their thing, and then provide a package of care back to the province that they come from." (C07) |
| **2. Uncertain and variable clinical Pathway** | No standardized pathway to motility specialists | "I don't think we have a great framework for how we approach them... I'm sure practice varies hugely between, even within, departments, let alone between departments and between gastroenterologists." (C09)<br>"It's a very variable [pathway] and I think the lack of consistency is a poor message to our patients who are already feeling rubbish... I do think a more universal approach is a really good idea." (C04) |
|  | Time to diagnosis varies significantly | "If someone is very classic then you can make the diagnosis on what's right then and there... If it's less typical, it may take a bit longer." (C05) |
| **3. Nuance of investigations** | Extensive Testing | "Usually they have had every test that could be done, has been done by the time they get to me... He [referring physician] doesn't like to leave any stones unturned." (C07) |
|  | Benefits of testing | "I think often the patient's symptoms don't always improve, but it's about being able to reassure the patient that we've actually done the right tests." (C08) |
|  | Inadequacies with testing | "Let's say they come and they've had no investigation, then it's going to be months to get all the investigations done." (C04)<br>"If it [testing] predicted a response or helped to define a subgroup of patients then I think that sort of test would be useful." (C06) |
| **4. Difficult to therapeutically manage** | Inadequate treatment options | "... severe gastroparesis or severe chronic unexplained nausea and vomiting is very difficult to treat." (C02)<br>"I've had a few [patients] who have gone off and had things like gastric electrical stimulators put in, but I have to say that the hit rate of those improving any symptom is you know about zero percent." (C06) |
|  | Trial and error therapy | "We just had to sort of trial and error you know... The patients get frustrated with a constant trial of medication after medication without sort of any sort of guarantee that we're actually trying the right thing." (C08) |
|  | Need for a multimodal approach | "I think that if it's been determined that patients are not going to benefit from, or unlikely to benefit from, medication or interventional therapies, then I'd be very happy for psychosocial support services to take over and hopefully give patient benefit." (C07)<br>"I have a few dietitians, who I'll refer people to. It's partly for symptom control and partly for maintenance of nutrition." (C06) |
| **5. Barriers to the therapeutic relationship** | Patient expectation of diagnosis | "They've [patients] seen a whole bunch of other specialists and you're sort of like the last port of call, and you're expected to come up with this diagnosis and treatment plan." (C05)<br>"Often we find that patients are looking for a label of some sort and I need to be careful about labeling somebody... until we have the [test] results." (C02) |
|  | Difficulty managing therapeutic relationship | "They [patients] haven't had good experiences at the point that you meet them. So there's a lot of unpicking that needs to be done." (C04)<br>"I think sometimes broaching some of those issues around eating disorders and psychological issues can sometimes compromise your doctor-patient relationship." (C07) |
|  | Clinician frustration | "I find that they require a lot of work... It's very frequent follow ups." (C05)<br>"It really causes people a lot of anxiety to keep seeing the same [patient] names coming up." (C07)<br>"That is reversed stigma... a lot of clinicians, it seems, shudder at the thought of gastroparesis." (C04) |

#### Theme 1: Clinical complexity

The first key clinician theme explored the clinical complexity of these disorders. These were seen as a *complex set of disorders* with one clinician describing them as “a puzzle” (C08). Clinicians admitted a lack of knowledge of these disorders and noted there was little research to help guide their clinical management. Much of the complexity arose from the multifactorial, largely unknown etiology underlying these disorders, alongside the overlapping, heterogeneous, and refractory symptom presentation. Overall, the complexity of these disorders resulted in *difficulty of diagnosis*. The current diagnostic approaches (e.g. the Rome Criteria and gastric emptying tests) were seen as vague, unreliable, and unable to capture the nuance of the symptom presentations. Instead, clinicians relied more heavily on patient history and clinical experience. This complexity resulted in an overarching desire for a *multidisciplinary collaborative approach*. Clinicians suggested the development of a centralized, multidisciplinary service to avoid delays and long wait times associated with referrals, and to allow for different specialists to work together to develop a thorough care plan for patients.

#### Theme 2: Uncertain and variable clinical pathway

The second key clinician theme explored the uncertainty and variability in the clinical care pathway. Clinicians discussed how there was *no standardized pathway to motility specialists* and how this impacted patient care. This lack of standardization often led to delays in referral from primary care and resulted in stagnation. Clinicians noted regional and international variations in the pathway for these patients, as well as differences in the approaches of individual clinicians. The multitude of differences meant that each patient appeared to proceed to the clinician through a different pathway. This resulted in an overarching desire for a more standardized and universal pathway. Because of the lack of standardization, the *time to diagnosis for these patients varied significantly*. Most patients already had symptoms for years before they were referred to a specialist and diagnosis further took months to years from the point of referral.

#### Theme 3: Nuance of investigations

The third clinician theme explored the nuance behind diagnostic testing and investigations. Clinicians described the *extensive testing* these patients underwent; however, which tests were completed was variable and dependent on the individual healthcare provider. The clinicians further discussed the *benefits of testing* including how test results helped them to exclude possible diagnoses. Testing helped inform treatment decisions and provided reassurance to patients that clinicians were making an effort towards diagnosis. However, many *inadequacies with testing* were also mentioned. For example, the accessibility of tests was variable and there were often long wait times. Tests were often low yield and clinicians reported a lack of reliability and standardization, especially for gastric emptying tests, which lowered their confidence in the results. Due to these concerns, clinicians iterated a need for better testing that could discriminate between different functional disorders and guide treatment decisions with more certainty.

#### Theme 4: Difficult to therapeutically manage

The fourth key clinician theme emphasized the difficulty in therapeutically managing these conditions. Clinicians highlighted the *inadequacies of treatment options* available and detailed their concerns about the lack of a “magic elixir” (C04) which was often desired or expected by patients. Management was usually done via *trial and error therapy*, where clinicians would trial different treatments to see what each patient responded to. The clinicians highlighted the *need for a multimodal approach* to manage these disorders that didn’t just rely on medications. The importance of psychological support was discussed at length; however, clinicians noted the challenge in accessing these services and wanted more psychological support to be available. Dietary input was also seen as important to reduce symptom burden and prevent malnutrition.

#### Theme 5: Barriers to the therapeutic relationship

The last key clinician theme concerned the various barriers to the therapeutic relationship. Firstly, it was highlighted that *patients usually had an expectation of diagnosis* to validate their symptoms. Some patients seemed to come to their appointments with a predetermined diagnosis and appeared to push the clinician to confirm this (self)-diagnosis. This put pressure on clinicians to manage patient expectations and resulted in increased caution regarding diagnosis. Clinicians reported concern about giving a diagnosis too early when there was not sufficient supporting evidence for it.

Alongside this, clinicians further discussed *the difficulty they had managing the therapeutic relationship*. Many patients had experienced previous strained clinical encounters and therefore appeared to enter the relationship with negative expectations. Reassuring these patients was seen as a challenge, especially when many did not receive a clear diagnosis or effective treatment. There was fear that this would be perceived by patients as mismanagement or lack of effort by clinicians. Lastly, this theme highlighted a degree of *clinician frustration* with the management of these disorders. Diagnosing and managing these disorders was seen as both time and resource intensive. There was an overarching feeling of frustration and anxiety with the lack of clinical improvement, despite the effort of the clinicians. These factors, alongside the complexity and challenge of these disorders, resulted in a degree of stigma and avoidance of these disorders for some clinicians.

## Discussion

The present study utilized semi-structured interviews to gain insight into the experiences of the current NVS and FD clinical care pathway from both a patient and gastroenterologist perspective. Themes identified within this sample suggest that both groups were dissatisfied with the current clinical care pathway, due to the perceived inadequacies in both diagnosis and treatment of these disorders. Based on this, there was a desire for a more standardized and holistic approach.

Overall, patients and clinicians agreed that the current clinical care pathway is complex, uncertain, and highly variable across gastroenterologists, departments, hospitals, regions, and nations. Obtaining a diagnosis and effective symptom management plan was seen as time-intensive and inefficient, requiring repeated contact points with the health system, exhaustive testing, and extensive trial and error therapy. These findings conform with existing international literature ^17,21,35^. The consistency of the present findings highlights that both groups are frustrated with the current clinical care pathway and are therefore calling for improvements. Patients and clinicians both desired more useful diagnostic testing options. This would help reduce the impact of the current trial and error model of treatment, which is solely based on symptomatology and is burdensome for both patients and the healthcare system ^17,22,36^. Testing options that utilize actionable biomarkers could help standardize the clinical care pathway, enable clinicians to better communicate the condition with their patients, and increase consistency of care ^1,37^. In turn, this could improve the therapeutic relationship and ultimately patient and clinician satisfaction ^38^.

There was also a call from both patients and clinicians for a multidisciplinary approach, which integrates gastroenterologists, dieticians, and psychologists into the management of these conditions. While patients appreciated the unique contributions of each healthcare provider within their broader care team, more cohesive coordination and communication were desired. Clinicians echoed this, noting that although it would be beneficial to provide a holistic model of care, the current system and lack of staff resources often made these services difficult to access. These findings are supported by Bray and colleagues ^39^ who demonstrated the efficacy of a multidisciplinary, integrated treatment approach to managing patients with FD and irritable bowel syndrome. However, it is noted that caution is needed to ensure that the care pathway optimizes cohesive collaboration across different healthcare providers and serves a clinically diverse population ^40^.

Although patients and clinicians agreed on the complex nature of the clinical care pathway and areas for improvement, the perceived underlying reasons for the diagnostic and management challenges for these conditions differed. This incongruence reflects a point of disconnect between patients and clinicians, which further complicates the therapeutic relationship. For example, while clinicians discussed how the lengthy diagnostic process was partly due to the lack of knowledge and research around these conditions, the need to be cautious with labeling patients, and a fear of misdiagnosis, none of the patients mentioned this. Instead, the long diagnostic process was perceived by patients as a lack of interest, empathy, and proactivity by clinicians. This was reported for both diagnosis and management. This mismatch of perspectives reflects the miscommunication noted and aligns with existing qualitative research, where a lack of communication was seen as a barrier to effective care ^29,41^.

The mismatch of perspectives between patients and clinicians, combined with the lack of effective treatments and exhaustive testing, was reported as frustrating by both patients and clinicians. The lack of clinical improvement, despite significant clinical input, was a source of frustration for clinicians and created stigma towards managing functional conditions, which further complicated care. On the contrary, patient participants perceived the absence of clinical improvement and poor communication as their clinicians not taking their concerns seriously. This is consistent with the patients’ reported need to advocate for care, a factor which clinicians perceived added an extra layer of complexity in an already complicated dynamic. This appeared to contribute to further deterioration of an already fragile patient-clinician therapeutic relationship. Previous research supports the significance of an effective therapeutic relationship between patients and clinicians when managing chronic health conditions ^38,41^. Specifically within FGID patients, a lack of perceived empathy and communication from gastroenterologists have been found to significantly reduce patient satisfaction with care ^37^. Therefore, it is important to improve communication between patients and clinicians to enable a positive and constructive therapeutic relationship.

While data saturation was achieved in this study, some limitations are worth noting. Despite an extended recruitment period, the patient sample was predominantly Caucasian and 100% female; which may explain why referrals to and admissions under the gynecology service were identified as a common issue. Although NVS and FD are more prevalent in females ^6^, future research would benefit from exploring the experiences of male patients and those of ethnic minorities. It is also notable that although an international clinician sample was recruited, the patient sample was based solely in New Zealand, limiting the patient themes to the New Zealand clinical care pathway. However, it is reasonable to assume that the current themes are likely to be representative of those countries with similar healthcare systems, such as the United Kingdom ^41,42^.

Given the call for a streamlined multidisciplinary approach, future research could benefit from the inclusion of a broader range of healthcare providers, including, dieticians, nurses, general practitioners (GPs), pharmacists, health psychologists, and psychiatrists, who are also involved in the management of NVS and FD. Understanding the perspectives of these providers may aid the conceptualization of an ideal multidisciplinary model of care.

## Conclusions

The current study provides insight into the lived experiences of patients with NVS and FD and the gastroenterologists who see and treat these patients. Both patients and clinicians experienced long, convoluted clinical care pathways, with many ongoing negative impacts. Recommendations from patients and clinicians included the development of more useful diagnostic tests, standardization of the clinical care pathway, and the adoption of an integrated, multidisciplinary approach to diagnosis and treatment. Further research should aim to better understand the costs of the current clinical care pathway and therefore understand the impacts of the NVS and FD clinical care pathway on the health system as a whole.

## Acknowledgements and Funding

The authors would like to thank the patients and clinicians who took part in this study. We thank Daniel Carson, Nikita Karulkar, and Izzy Goddard for their technical assistance. This study was supported by the Health Research Council of New Zealand.

## Author Contributions

GS, ML, EB, AAG, CNA, CD, GOG, SC, CK were involved in study conception and design. CK and GS were involved in data collection. GS, ML, EB, AAG, CNA, GOG, SC, CK were involved in the data analysis and interpretation. All authors contributed to the drafting, critical revisions, and final approval of the manuscript.

## Data Availability

Statement The data that support the findings of this study are available from the corresponding author [CK], upon reasonable request.

